# Immunity to non-dengue flaviviruses impacts dengue virus IgG ELISA specificity in Cambodia

**DOI:** 10.1101/2023.11.17.23298701

**Authors:** Camila Odio, Christina Yek, Chloe M. Hasund, Somnang Man, Piseth Ly, Sreynik Nhek, Sophana Chea, Chanthap Lon, Charlie Voirin, Rekol Huy, Rithea Leang, Chea Huch, L. Fabiano Oliveira, Jessica E. Manning, Leah C. Katzelnick

**Affiliations:** Viral Epidemiology and Immunity Unit, Laboratory of Infectious Diseases, National Institute of Allergy and Infectious Diseases, National Institutes of Health, Bethesda, MD 20892, USA; International Center of Excellence in Research, National Institute of Allergy and Infectious Diseases, National Institutes of Health, Phnom Penh, Cambodia; Laboratory of Malaria and Vector Research, National Institute of Allergy and Infectious Diseases, National Institutes of Health, Bethesda, MD 20892; National Center for Parasitology, Entomology, and Malaria Control, Ministry of Health, Phnom Penh, Cambodia

**Author notes:** **Corresponding authors:** Leah Katzelnick PhD MPH, Chief, Viral Epidemiology and Immunity Unit, National Institutes of Allergy and Infectious Diseases, National Institutes of Health, Building 33, Room 3W10A, 33 North Drive, Bethesda, MD 20892-3203, Jessica Manning MD MSc, Director, International Center of Excellence in Research CAMBODIA, National Institutes of Allergy and Infectious Diseases, National Institutes of Health. Authors made equal contributions to this manuscript (Christina Yek, Chloe M. Hasund).

## Abstract

Seroprevalence studies are the gold standard for disease surveillance, and serology was used to determine eligibility for the first licensed dengue vaccine. However, expanding flavivirus endemicity, co-circulation, and vaccination complicate serology results. Among 713 healthy Cambodian children, a commonly used indirect dengue virus IgG ELISA (PanBio) had a lower specificity than previously reported (94% vs. 100%). Of those with false positive PanBio results, 46% had detectable neutralizing antibodies against other flaviviruses, with the highest frequency against West Nile virus (WNV). Immunity to non-dengue flaviviruses can impact dengue surveillance and potentially pre-vaccine screening efforts.

## Background

The genus *Orthoflavivirus* includes multiple pathogenic mosquito-borne viruses including dengue viruses 1-4 (DENV1-4), Japanese encephalitis virus (JEV), West Nile virus (WNV), Zika virus (ZIKV) and yellow fever virus (YFV) [1]. With expanding vector habitats, known flaviviruses are rising in global incidence, and novel flaviviruses are emerging [2, 3]. These flaviviruses commonly co-circulate, and the antibodies induced by one exposure may cross-react with others in the genus [4]. Additionally, affected areas use vaccines to protect against JEV and YFV, further complicating serology. As flaviviruses expand their range and vaccination increases, differentiating true exposure from cross-reactivity is not only difficult, but also increasingly important to guide diagnostic, preventative, and therapeutic measures.

Accurately characterizing population level immunity to DENV is important for current dengue vaccination efforts. Dengvaxia (Sanofi Pasteur), the first widely approved dengue vaccine, was originally recommended in 2016 by the World Health Organization (WHO) Strategic Advisory Group of Experts (SAGE) panel for use in areas with ≥70% DENV seroprevalence in children age 9 and older, with seroprevalence most often measured using common IgG ELISAs [5]. When it was later shown that Dengvaxia increases the risk of severe disease in DENV-naïve individuals, the WHO recommended use of highly specific individual-level testing of DENV immunity to confirm vaccine eligibility, as well as use in high-risk populations in endemic areas [6][7]. The World Health Organization has since recommended that the second licensed dengue vaccine QDENGA (Takeda) be introduced to children aged 6 to 16 years, also in highly endemic areas [8]. Given the lack of observed vaccine-induced protection against DENV3 in seronegative individuals and the unknown protection against DENV4 [9], a strategy based on population-level estimates of endemicity has the potential to increase disease risk for seronegative individuals.

Population-level serosurveys for DENV conducted for surveillance purposes generally use commercial IgG ELISAs. The plaque reduction neutralization test (PRNT), which measures neutralizing antibodies (nAbs) to DENV, is considered the gold standard for evaluating specificity but requires intensive and specialized labor. The PanBio indirect DENV IgG ELISA (Abbott, Brisbane, QLD, Australia) is one of the most commonly used assays for measuring DENV immunity, and the manufacturer reports 100% specificity based on 108 DENV-naïve individuals from endemic areas [10]. Separate work demonstrated 99% specificity of this ELISA using DENV1-4 PRNT_50_≥10 as the standard for DENV immunity in a cohort of 534 individuals from both the USA and dengue-endemic regions before 2016 [11]. However, the PanBio indirect DENV IgG ELISA yields higher false positivity rates when evaluated with individuals positive to other flaviviruses, including those who had received an inactivated JEV vaccine (3%), or had immunity against ZIKV (34%) or WNV (51%) [11]. The degree to which immunity to other flaviviruses affects DENV serosurveys is dependent on the site, and a major challenge is that the extent of circulation of other flaviviruses is often unknown. For instance, an observational study of children aged 9-14 years in the Philippines in 2017 used a PRNT_70_≥40 as the indicator of DENV immunity and reported a relatively low ELISA specificity, of 93.4%. Of the false positive samples, 64% had nAb against ZIKV or JEV, in a region where ZIKV was not thought to be widespread. Thus, although the PanBio ELISA has a reportedly high specificity, this number may vary with flavivirus cross-reactivity and expanding co-circulation or vaccination.

Here, we examine the performance of the PanBio ELISA in young children in Cambodia, a highly dengue-endemic area. ZIKV was recently found to co-circulate in the area, and JEV is endemic. JEV vaccination campaigns with a live-attenuated JEV vaccine SA14-14-2 started around 2014, and WNV nAb have been identified in birds but not humans [12-15].

## Methods

The study protocol was approved by the institutional review boards at the US National Institutes of Health and the National Ethics Committee on Human Research in Cambodia. The guardians of all pediatric participants provided signed informed consent to participate in the study. Between July and August of 2018, 771 children aged 2-9 years living in Kampong Speu, Cambodia were enrolled in a prospective longitudinal cohort (NCT03534245) [16]. At entry, the PanBio indirect DENV IgG ELISA was performed on sera from 770 individuals. For the 273 participants with ELISA values >1.1 (defined as DENV positive by the manufacturer), PRNTs were performed using clinical isolates of DENV1-4 [17]. All PRNTs were performed on Vero cells, as described previously, starting at a serum dilution of 1:5 followed by the addition of an equal volume of virus. Thus, the lower limit of detection was a dilution of 1:10 [18]. The nAb titer was defined as the reciprocal of the calculated dilution wherein virus infectivity was reduced by 50% (PRNT_50_). PRNT_50_≥1:10 (reported as PRNT_50_≥10) against any DENV serotype was considered immune to that serotype. To assess for non-dengue flavivirus nAbs, PRNTs were performed using the following strains: ZIKV-PARAIBA/2015, a chimeric vaccine candidate, WNV/DEN4Δ30 [19], and JEV vaccine strain SA14-14-2 (Figure 1A-B). Consistent with prior classifications, seropositivity to any non-dengue flavivirus was defined as PRNT_50_≥10 [20-22]. Statistical comparisons were done in RStudio for macOS (2022.07.1, Build 554) using tidyverse and gtsummary packages. Heatmaps were generated in PRISM (v.9 for macOS).

**Figure 1.**
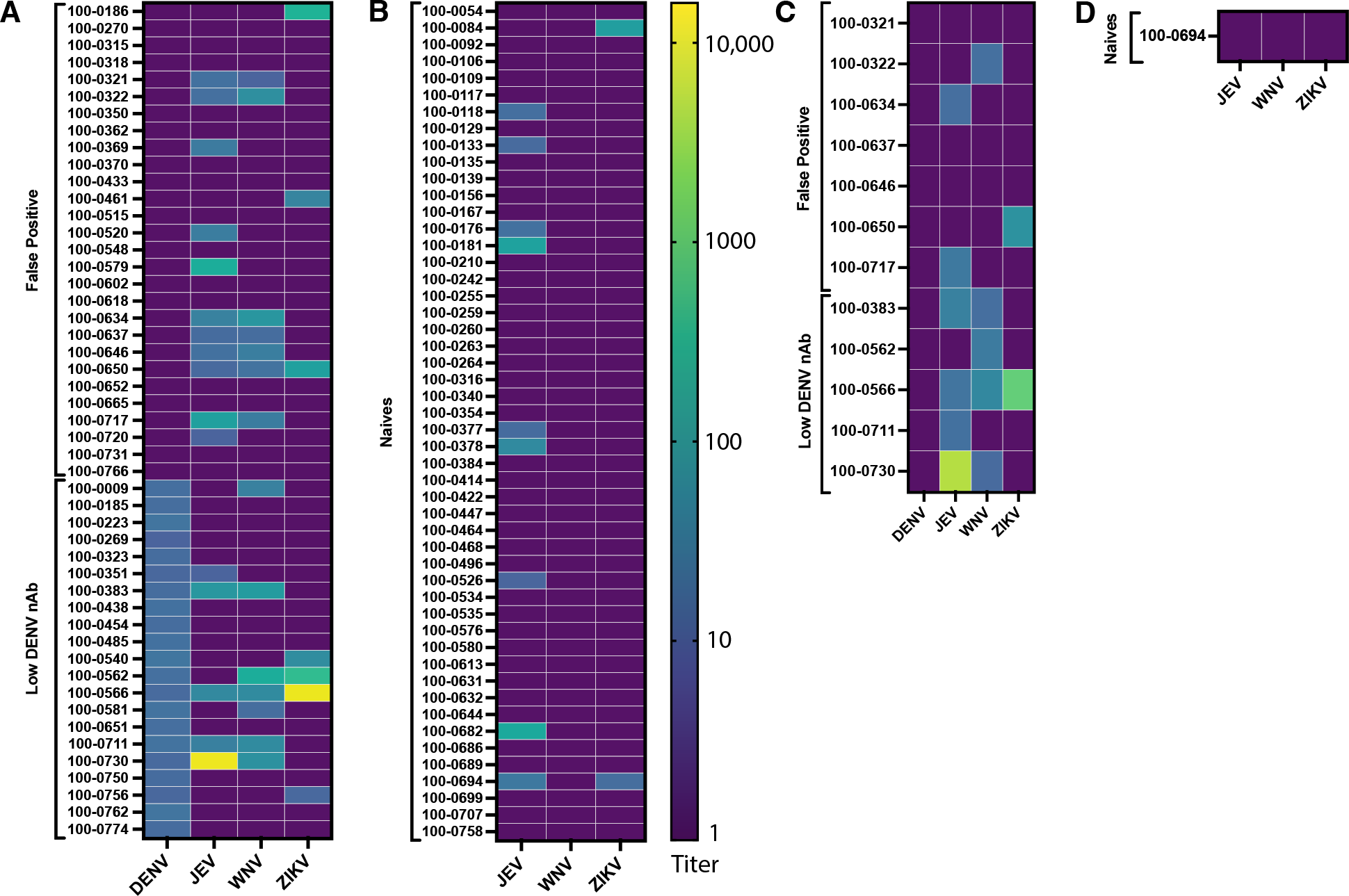
Neutralizing antibody titers against DENV, JEV, WNV, ZIKV in individuals who were false positive (ELISA>1.1, DENV PRNT_50_<10), naïve (ELISA<0.2), or had low DENV nAb (ELISA>1.1, DENV PRNT_50_ of 10-20) as measured by PRNT_50_ (A-B) and PRNT_90_ titers (C-D). PRNT_90_ titers were only measured in those with nAb against ≥2 non-dengue flaviviruses.

## Results

Of the 770 individuals, n=440 had ELISA<0.2 and were considered DENV naïve per work confirming a strong correlation between ELISA<0.2 and negative PRNT [23]. Fifty-seven individuals were excluded: 44 had ELISA values between 0.2-0.9 without a confirmatory PRNT and 13 had ELISA values between 0.9-1.1, which the manufacturer considers equivocal immunity. Of the 273 individuals with ELISA>1.1, n=28 were DENV negative by PRNT (‘false positive’), n=245 were DENV positive (‘immune’), and there were no false negatives, resulting in a 100% sensitivity and 94% specificity of the ELISA assay. Comparison of the naïve, false positive, and immune groups revealed that both the mean age and ELISA value of the false positive group fell between those of the naïve and immune groups (p<0.001, Table 1A). Over the two years of surveillance, there were few PCR-confirmed cases of symptomatic dengue (n=46) with no differences in frequency among groups (p=0.7).

**Table 1.** Characteristics and immune profiles of individuals who were naïve (ELISA<0.2), false positive (ELISA>1.1, PRNT_50_<10), or DENV immune (ELISA>1.1, PRNT_50_≥10). The presence of neutralizing antibodies against non-dengue flaviviruses were compared in the naïve and false positive groups and between the false positive and the low DENV immune group (ELISA>1.1, PRNT_50_ 10-20).

| Characteristics | Naïve<br>(n=440)* | False positive<br>(n=28) | Immune (n=245) | p-value<sup>†</sup> |
| --- | --- | --- | --- | --- |
| Age | 4.9 (2.0) | 6.0 (2.2) | 6.8 (2.2) | <0.001 |
| Sex |  |  |  | 0.2 |
| Female | 230 (52) | 10 (36) | 126 (51) |  |
| Male | 210 (48) | 18 (64) | 119 (49) |  |
| ELISA IgG value | 0.03 (0.04) | 2.00 (0.72) | 2.92 (0.84) | <0.001 |
| Dengue case |  |  |  | 0.7 |
| No dengue | 321 (91) | 22 (92) | 156 (93) |  |
| Symptomatic<br>dengue | 32 (9) | 2 (8) | 12 (7) |  |
| Unknown<sup>‡</sup> | 87 | 4 | 77 |  |

B. Immunity subset analysis
| Immunity<br>profile | Naïve<br>(n=50) | p-value<sup>§</sup> | False positive<br>(n=28) | p-value<sup>¶</sup> | Low DENV nAb<br>(n=21) |
| --- | --- | --- | --- | --- | --- |
| JEV nAb |  | 0.058 |  | 0.4 |  |
| Negative | 41 (82) |  | 17 (61) |  | 16 (76) |
| Positive | 9 (18) |  | 11 (39) |  | 5 (24) |
| ZIKV nAb |  | 0.3 |  | 0.4 |  |
| Negative | 48 (96) |  | 25 (89) |  | 17 (81) |
| Positive | 2 (4) |  | 3 (11) |  | 4 (19) |
| WNV nAb |  | <0.001 |  | 0.5 |  |
| Negative | 50 (100) |  | 21 (75) |  | 14 (67) |
| Positive | 0 (0) |  | 7 (25) |  | 7 (33) |
| ≥1 Flavivirus<br>positive | 10 (20) | 0.020 | 13 (46) | >0.9 | 10 (48) |
| ≥2 Flavivirus<br>positive | 1 (2) | 0.003 | 7 (25) | >0.9 | 5 (24) |

\*Mean (SD) or no. (%)
†One-way ANOVA, Pearson's Chi-squared test; Fisher's exact test
‡Unknown indicates that participant was lost to follow-up prior to final study visit at 24 months
§Fisher's exact test comparing naïve vs. false positive group.
¶Fisher's exact test comparing false positive group vs. low DENV nAb group.
Dengue virus, DENV; Japanese encephalitis virus, JEV; neutralizing antibodies, nAb; West Nile
virus, WNV; Zika virus, ZIKV

We hypothesized that the discordance between the DENV ELISA IgG and PRNT results could be due to cross-reactivitiy against non-dengue flaviviruses. To test this, we compared the frequencies of nAbs against JEV, ZIKV, and WNV in the false positive individuals versus n=50 randomly chosen naïve individuals (Figure 1A-B). Consistent with prior classifications, seropositivity was defined as PRNT_50_≥10 [20-22]. Overall, 46% of the false positive group had PRNT_50_≥10 against ≥1 other flavivirus versus 20% of the naive group (p=0.020, Table 1B). Although the false positive group had higher percentages of individuals with positive JEV and ZIKV nAbs, only WNV nAbs were significantly more common than in the naïve group (0% vs. 25%, p<0.001). To further assess these trends, we tested n=21 individuals with DENV ELISA>1.1 and DENV PRNT between 10-20 (Figure 1). This group has low DENV nAbs and has been considered DENV negative in other work [16, 23]. When compared to the false positive group, the low DENV nAb group had similar frequencies of JEV, ZIKV, and WNV nAb (p≥0.4 for all three nAbs, Table 1B). Thus, immunity to other flaviviruses may contribute to high ELISA values in individuals with undetectable and low DENV nAbs.

To help identify the primary exposure in the 13 individuals with PRNT_50_≥10 against ≥2 flaviviruses, PRNT_90_ titers were calculated [4]. Of these, n=4 had PRNT_90_<10 against all 4 flaviviruses, and n=3 had PRNT_90_≥10 to multiple flaviviruses: n=2 to JEV and WNV, n=1 to JEV, WNV, and ZIKV (Figure 1C-D). Six individuals had PRNT_90_≥10 against only 1 flavivirus: n=1 to ZIKV, n=3 to JEV, and n=2 to WNV. Of the two individuals with PRNT_90_≥10 against WNV only, one had received a JEV vaccine and had a PRNT_50_≥10 against JEV and one had PRNT_50_≥10 against ZIKV. While the PRNT_90_≥10 against WNV only suggests a primary WNV infection, it is possible that JEV vaccination and ZIKV primary exposure induced highly cross-reactive WNV nAb [24]. There were also two individuals with low DENV nAb and PRNT_50_≥10 against WNV only. Although these individuals did not have PRNT_90_≥10 against DENV or WNV, one had WNV titer that was 2-fold higher than DENV titer. Again, this finding is suggestive of primary WNV infection, but cross-reactivity after DENV exposure cannot be ruled out. Regardless of the WNV nAb source, this immunity was common and likely central to the decreased ELISA specificity observed.

## Discussion

We found that the PanBio DENV indirect IgG ELISA has a lower specificity than reported in prior studies. This discrepancy may be partially explained by the assay’s detection of WNV nAb with potential contributions by ZIKV and JEV nAb. Notably, JEV vaccination can induce WNV cross-reactivity [25], and it is possible that the WNV nAb were induced by JEV vaccination or infection. Alternatively, the WNV nAbs may represent true WNV exposure, underlining the need for ongoing vigilance for WNV circulation in humans in Cambodia. Additionally, half of the false positive results remained unexplained, potentially due to waning immunity or infection by unidentified flaviviruses.

Clinicians, investigators, and public health authorities should be aware that expanding flavivirus co-circulation and vaccination could increasingly impact serology results. Serosurveys conducted for vaccination campaigns to identify populations where dengue is endemic may overestimate dengue burden as a result of false positivity due to infection or vaccination with other flaviviruses. Such population-based strategies are of particular concern when identifying target populations for dengue vaccines where safety in DENV seronegative individuals has not yet been confirmed. Adverse events in these individuals could greatly impact vaccine trust and uptake, as occurred with Dengvaxia [26]. For vaccines that are known to be unsafe in seronegative individuals like Dengvaxia, pre-vaccination screening is required to determine vaccine eligibility, which allows individuals to make informed decisions about their own vaccine risk and benefit. It is recommended that past infection be confirmed either by virological assay or by two specific serological assays, such as the anti-DENV1-4 NS1 ELISA IgG and a IgG rapid test [7]. Evaluation of false positivity due to infection with other emerging flaviviruses is critical to ensuring the safety of this screening approach.

Overall, our study demonstrates that the PanBio IgG ELISA and even PRNT results should be interpreted with caution in areas with flavivirus co-circulation and vaccines, and multiple tests may be required to confirm DENV seroprevalence.

## Data Availability

All data produced in the present study are available upon reasonable request to the authors.

## Acknowledgements

The authors would like to thank Patrick Mpingabo for his support establishing ELISA assays to evaluate samples for antibodies against the nonstructural-1 proteins of JEV, ZIKV, and WNV. These assays were not ultimately used in the manuscript. We would also like to thank Kelsey Lowman for her support completing the WNV and JEV PRNT assays.

## Conflict of Interest

The authors declare no conflict of interest.

This work was presented as a poster at the American Society of Tropical Medicine and Hygeine annual conference in October 2023.

## References

1. Postler TS, Beer M, Blitvich BJ, et al. Renaming of the genus Flavivirus to Orthoflavivirus and extension of binomial species names within the family Flaviviridae. Archives of Virology 2023; 168:224.

2. Du M, Jing W, Liu M, Liu J. The Global Trends and Regional Differences in Incidence of Dengue Infection from 1990 to 2019: An Analysis from the Global Burden of Disease Study 2019. Infectious Diseases and Therapy 2021; 10:1625–43.

3. Pierson TC, Diamond MS. The continued threat of emerging flaviviruses. Nature Microbiology 2020; 5:796–812.

4. Chan KR, Ismail AA, Thergarajan G, et al. Serological cross-reactivity among common flaviviruses. Front Cell Infect Microbiol 2022; 12:975398.

5. Secretariat SWGoDVaWHO. Background paper on dengue vaccines. Available at: https://terrance.who.int/mediacentre/data/sage/SAGE_Docs_Ppt_Apr2016/10_session_dengue/Apr2016_session10_dengue_vaccines.pdf. Accessed 11/17/2023.

6. WHO. Dengue vaccine: WHO position paper. Weekly epidemiological record 2018; 93:457–76.

7. Prevention CfDCa. Laboratory Testing Requirements for Vaccination with Dengvaxia Dengue Vaccine. Available at: https://www.cdc.gov/dengue/vaccine/hcp/testing.html. Accessed October 31 2023.

8. Takeda. Takeda’s Dengue Vaccine Recommended by World Health Organization Advisory Group for Introduction in High Dengue Burden and Transmission Areas in Children Ages Six to 16 Years. Available at: https://www.takeda.com/newsroom/newsreleases/2023/Takeda-Dengue-Vaccine-Recommended-by-World-Health-Organization-Advisory-Group-for-Introduction-in-High-Dengue-Burden-and-Transmission-Areas-in-Children-Ages-Six-to-16-Years/. Accessed October 31 2023.

9. Rivera L, Biswal S, Sáez-Llorens X, et al. Three-year Efficacy and Safety of Takeda’s Dengue Vaccine Candidate (TAK-003). Clinical Infectious Diseases 2021.

10. Abbott. Panbio™ DENGUE IgG CAPTURE ELISA. Available at: https://www.globalpointofcare.abbott/au/en/product-details/panbio-dengue-igg-capture-elisa.html. Accessed August 29 2023.

11. Bonaparte M, Zheng L, Garg S, et al. Evaluation of rapid diagnostic tests and conventional enzyme-linked immunosorbent assays to determine prior dengue infection. J Travel Med 2019; 26.

12. Yek C, Li Y, Pacheco AR, et al. National dengue surveillance, Cambodia 2002-2020. Bull World Health Organ 2023; 101:605–16.

13. Duong V, Ong S, Leang R, et al. Low Circulation of Zika Virus, Cambodia, 2007-2016. Emerg Infect Dis 2017; 23:296–9.

14. Horwood PF, Duong V, Laurent D, et al. Aetiology of acute meningoencephalitis in Cambodian children, 2010-2013. Emerg Microbes Infect 2017; 6:e35.

15. Auerswald H, Ruget AS, Ladreyt H, et al. Serological Evidence for Japanese Encephalitis and West Nile Virus Infections in Domestic Birds in Cambodia. Front Vet Sci 2020; 7:15.

16. Manning JE, Chea S, Parker DM, et al. Development of Inapparent Dengue Associated With Increased Antibody Levels to Aedes aegypti Salivary Proteins: A Longitudinal Dengue Cohort in Cambodia. The Journal of Infectious Diseases 2021.

17. Durbin AP, Karron RA, Sun W, et al. Attenuation and immunogenicity in humans of a live dengue virus type-4 vaccine candidate with a 30 nucleotide deletion in its 3’-untranslated region. Am J Trop Med Hyg 2001; 65:405–13.

18. Timiryasova TM, Bonaparte MI, Luo P, Zedar R, Hu BT, Hildreth SW. Optimization and validation of a plaque reduction neutralization test for the detection of neutralizing antibodies to four serotypes of dengue virus used in support of dengue vaccine development. Am J Trop Med Hyg 2013; 88:962–70.

19. Pierce KK, Whitehead SS, Kirkpatrick BD, et al. A Live Attenuated Chimeric West Nile Virus Vaccine, rWN/DEN4Δ30, Is Well Tolerated and Immunogenic in Flavivirus-Naive Older Adult Volunteers. The Journal of Infectious Diseases 2016; 215:52–5.

20. Plotkin SA. Correlates of protection induced by vaccination. Clin Vaccine Immunol 2010; 17:1055–65.

21. Sornjai W, Jaratsittisin J, Auewarakul P, Wikan N, Smith DR. Analysis of Zika virus neutralizing antibodies in normal healthy Thais. Sci Rep 2018; 8:17193.

22. Kaiser JA, Barrett ADT. Twenty Years of Progress Toward West Nile Virus Vaccine Development. Viruses 2019; 11.

23. Lopez AL, Adams C, Ylade M, et al. Determining dengue virus serostatus by indirect IgG ELISA compared with focus reduction neutralisation test in children in Cebu, Philippines: a prospective population-based study. The Lancet Global Health 2021; 9:e44–e51.

24. Hou B, Chen H, Gao N, An J. Cross-Reactive Immunity among Five Medically Important Mosquito-Borne Flaviviruses Related to Human Diseases. Viruses 2022; 14:1213.

25. Mansfield KL, Horton DL, Johnson N, et al. Flavivirus-induced antibody cross-reactivity. J Gen Virol 2011; 92:2821–9.

26. Larson HJ, Hartigan-Go K, de Figueiredo A. Vaccine confidence plummets in the Philippines following dengue vaccine scare: why it matters to pandemic preparedness. Hum Vaccin Immunother 2019; 15:625–7.

